# Factors affecting the transmission of SARS-CoV-2 in school settings

**DOI:** 10.1101/2021.06.18.21259156

**Authors:** Haokun Yuan, Connor Reynolds, Sydney Ng, Wan Yang

**Author notes:** Correspondence to: Wan Yang, Department of Epidemiology, Mailman School of Public Health, Columbia University, 722 W 168th Street, Room 514, New York, NY 10032. Authors contributed equally (Haokun Yuan, Connor Reynolds, Sydney Ng).

## Abstract

**Background:** Several studies have reported SARS-CoV-2 outbreaks in schools, with a wide range of secondary attack rate (SAR; range: 0-100%). We aimed to examine key risk factors to better understand SARS-CoV-2 transmission in schools.

**Methods:** We collected records of 39 SARS-CoV-2 school outbreaks globally published through July 2021 and compiled information on hypothesized risk factors. We utilized the directed acyclic graph (DAG) to conceptualize risk mechanisms, used logistic regression to examine each risk-factor group, and further built multi-risk models.

**Results:** The best-fit model showed that the intensity of concurrent community transmission (adjusted odds ratio [aOR]: 1.2, 95% CI: 1.17 – 1.24, for each increase of 1 case per 10,000 persons per week), individualism (aOR: 1.72, 95% CI: 1.19 – 2.5, above vs. below the median) were associated higher risk, whereas preventive measures (aOR: 0.22, 95% CI: 0.17 – 0.29, distancing and masking vs. none) and higher population immunity (aOR: 0.28, 95% CI: 0.22 – 0.35) were associated with lower risk of SARS-CoV-2 transmission in schools. Compared to students in pre-schools, the aOR was 0.35 (95% CI: 0.23 – 0.54) for students in primary schools and 1.3 (95% CI: 0.9 – 1.88) for students in high schools.

**Conclusions:** Preventive measures in schools (e.g. social distancing and mask-wearing) and communal efforts to lower transmission and increase vaccination uptake (i.e. vaccine-induced population immunity) in the community should be taken to collectively reduce transmission and protect children in schools. Flexible reopening policies may be considered for different levels of schools given their risk differences.

## Introduction

Since the early stages of the COVID-19 pandemic, concerns have been raised about the impact of schools on community transmission and the well-being of students and staff, as well as the impact on the schedules of healthcare workers concerning childcare [1]. Out of an abundance of caution and fear that the SARS-CoV-2 virus would spread rapidly in schools much like influenza pandemics [2], countries globally decided to suspend in-person classes and begin online instruction. By April 2020, over 600 million students worldwide were affected by school closures in response to the COVID-19 pandemic [3]. In contrast to influenza pandemics where children are the key drivers of transmission, studies have indicated that children are likely less susceptible to SARS-CoV-2 infection, tend to experience less severe disease when infected, and likely have lower transmissibility [4, 5]. Given this new evidence, schools in many places have gradually reopened since the summer of 2020, while implementing varying level of preventive measures (e.g., mask wearing, distancing, limiting the number of students, rotating schedules, and viral testing) to reduce risk of transmission. Given these circumstances, the risk of SARS-CoV-2 outbreaks in school settings may differ substantially across space and time. Indeed, several studies have examined school outbreaks of COVID-19 and reported secondary attack rates (SAR) – i.e., the proportion of infected contacts of an index case out of all contacts of that index case [6] – ranging from 0 (i.e., no secondary infections) to 100% (i.e., infections among all contacts). However, this discrepancy is still not fully understood, and a better understanding can inform better preventive measures for future outbreaks not limited to COVID-19 or school settings.

To identify the main factors that determine the transmission of SARS-CoV-2 in schools and inform strategies to prevent future school outbreaks, here we examined the associations between SARS-CoV-2 SAR in children and various potential risk factors. We compiled data from relevant studies in the literature reporting SARS-CoV-2 SAR in schools and for related factors (e.g. incidence in the community and population immunity cumulated over time) and further used regression models to examine key risk factors of having high SAR in schools. Consistent with previous work, we found the risk varied by school level, with lower risk among primary school students than preschoolers and high schoolers. Accounting for school level, we found that implementation of preventive measures (distancing and mask wearing) in schools and higher population immunity were associated lower SAR in schools; in contrast, higher SARS-CoV-2 transmission in the community and higher level of individualism were associated with higher SAR in schools.

## Methods

### Data sources

Studies were searched for on the “Living Evidence for COVID-19” database [7], which retrieves articles from EMBASE via Ovid, PubMed, BioRxiv, and MedRxiv. Any article within this database was considered, from December 2019 up to July 28, 2021. The search terms used include “transmission AND (school OR schools)” or “transmission AND children”. When titles and abstracts were identified as being potentially relevant, the articles were read to determine if the outbreaks took place in a school setting and the number of infections and contacts were reported. In addition, we extracted 11 observations included in a systematic review of evidence regarding the ability of children to transmit SARS-CoV-2 in schools [8]. In total, 39 school outbreaks extracted from 26 articles were included in this analysis (see Fig 1).

**Figure 1.**
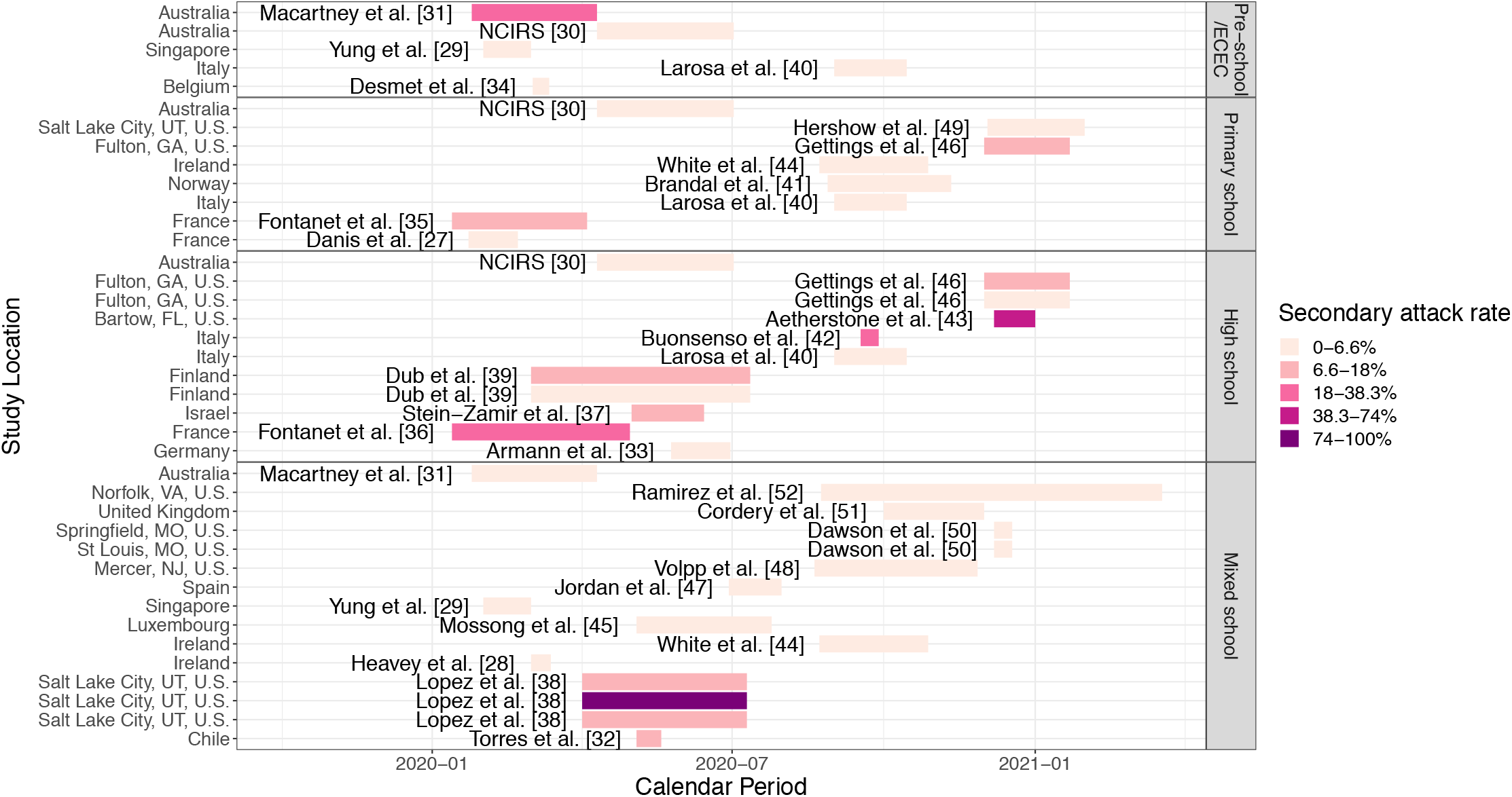
SARS-CoV-2 school outbreak studies included in the analysis [27-51]. Each colored bar represents an observed school outbreak; the school location is shown on the y-axis and study period is shown by the position and length of the bar (see calendar time on the x-axis); school type is shown in the panel title on the right; and reported secondary attack rate (SAR) is indicated by the color of the bar (see the legend).

Relevant data, as deemed by an initial conceptual analysis using the directed acyclic graph (DAG; see details below), were taken from the articles identified above. These included the time period of the study, study design, location, season, age of children, whether the study happened during a lockdown, type of school according to the International Standard Classification of Education [9], reported SARs, number of contacts of the index case, testing method (PCR vs. serology), level of surveillance (all contacts, some contacts, only symptomatic), and whether masks and social distancing were required. In addition, we compiled additional data for potential risk or confounding factors of SARS-CoV-2 transmission in schools for each identified study as detailed in the next section.

### Conceptual analysis and variable coding

The unit of analysis was individual study and in cases where several school types were covered in one study, the data were stratified by those school types. We first conducted a conceptual analysis using the DAG and identified nine key components that may affect SARS-CoV-2 transmission in schools (Fig 2). Below we describe each of the nine components, rationale for inclusion, and related variables examined.

**Figure 2.**
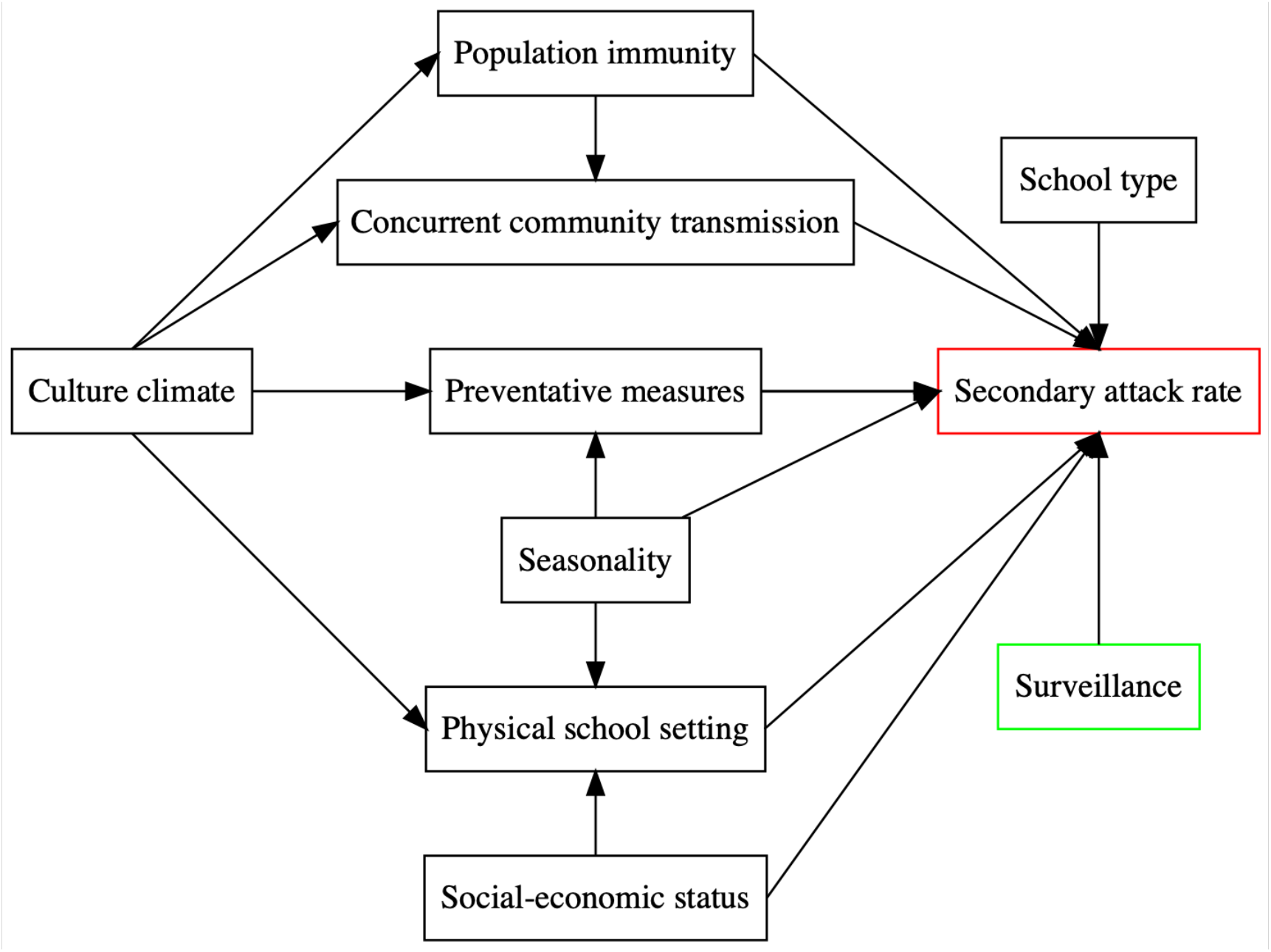
Directed acyclic graph (DAG) describing the relationship among variables. This DAG represents the meaningful relationships between the variables relevant to SARS-CoV-2 SAR among children in school settings and informs all further analyses. The outcome measurement, SAR, is presented in red while risk factors are in black and surveillance in green.

1. School types, based on studies indicating differential transmission risk among different age groups [10, 11]. Here, we examined this factor as a categorical variable including 4 levels, i.e. pre-school or early childhood education center (ECEC), primary school, high school, and mixed-level school. The first three levels were per reports in the included school studies. For studies that examined several types of school but did not report school type specific SARs, we assigned them to a “mixed-level school” category. For example, if a study gave the overall SAR combining a pre-school and a primary school, it was given the value “mixed-level school.” SARS-CoV-2 SAR among children in school settings is the number of infected contacts divided by the total number of contacts of the index cases at each school.
2. Physical school settings such as student density in the classroom and ventilation systems that may affect the intensity of school contact and clearance of air. As it is difficult to obtain information related to ventilation settings, here we included class size in our analysis based on the average number of students per classroom in each country, as reported by the Organisation for Economic Co-operation and Development (OECD) [12].
3. Preventive measures, which may reduce outbreak risk. Here we categorized this variable based on the implementation of mask-wearing and/or social distancing in schools, i.e., “No preventive measures” if neither measure was required, “Single preventive measure” if only one measure (i.e. distancing or masking) was required, and “Combined preventive measure” if both were required. Note that we were not able to test distancing and masking separately due to the small sample size of schools that required masking alone (n = 3).
4. Surveillance and/or testing policies implemented in schools. On the one hand, testing policies could affect the reported values of SAR; for instance, testing of all contacts regardless of symptoms may lead to identification of more infections including those asymptomatic and increase the numerator of SAR. On the other, frequent testing of all if combined with school closure may serve as a containment measure to reduce the risk of onward transmission and in turn reduce SAR. Here we thus included the reported testing practices for contacts in the school outbreak clusters as a categorical ordinal variable. Three types of testing were reported in the school studies, including testing only the symptomatic, both symptomatic and some asymptomatic, and all contacts. However, due to the small sample size in “only symptomatic” (n = 3), we dichotomized surveillance to testing “only symptomatic or some asymptomatic” and “all contacts” of an index case in each school cluster.
5. Seasonal changes such as humidity and temperature, which may affect the survival and transmission of SARS-CoV-2. Here, we used specific humidity (a measure of absolute humidity) to examine the potential impact from disease seasonality, as specific humidity and temperature are highly correlated. Specifically, ground surface temperature and relative humidity for each study location were extracted from the National Oceanic and Atmospheric Administration using the “rnoaa” package [13]. Daily mean specific humidity in g H20/kg air was then computed based on the meteorological data using formula introduced by Bolton [14], and further averaged over the corresponding study period.
6. Intensity of concurrent community transmission, which may increase the introduction of infections into schools. To examine its impact, we included two measures, i.e., the weekly COVID-19 case rate and weekly COVID-19-related death rate for the study area during the study period; both measures were computed using data from the John Hopkins Coronavirus Resource Center [15] and standardized by the corresponding population size (for non-US sites, country-level data were used; and for US sites, county-level data were used). In addition, testing and hence the case-ascertainment rate tended to increase over time, whereas infection-fatality risk tended to decrease over time due to, e.g., more timely diagnosis and improved medical treatments and management [16]. To account for such time-related impacts on the reported case rates and death rates, we included an additional variable (“calendar period”) whenever either measure is included in the model. We dichotomized the “calendar period” variable, based on whether a study was conducted before July 2020, i.e., roughly the end of the initial pandemic wave when both testing capacities and medical treatments were improved.
7. Prior population immunity in the community. Population immunity gained from prior infections or COVID-19 vaccination could lower population susceptibility and hence the risk of SARS-CoV-2 in the community. As most school outbreaks included here occurred prior to the rollout of mass-vaccination, population immunity at those times would mostly come from natural infections (see Fig 1 for the timeline of each study, vs. earliest vaccination rollout for the general population round spring 2021). Thus, here we used the cumulative COVID-19 case rate (up to the mid-point of the corresponding study period) as a proxy to account for prior population immunity.
8. Cultural climates, which “represent independent preferences for one state of affairs over another that distinguish countries (rather than individuals) from each other” [17] and may reflect the collective risk tendency of a population. The Hofstede’s cultural dimensions theory [17] included 6 related measures including individualism, masculinity, uncertainty avoidance, long term orientation, and indulgence. In particular, individualism is defined as the degree of interdependence of society maintains among its members. We reasoned that the individualism measure would be most relevant to the level of compliancy to public health interventions and in turn the risk of SARS-CoV-2 transmission. Thus, here we included individualism in our analysis and dichotomized the reported values for each country [18]. Among all study sites included here, the median of individualism scores was 76; thus, we coded those with a score >76 as “Higher individualism” those with a score ≤76 as “Lower individualism”.
9. Indicators of socioeconomic status such as national income that reflect a country’s ability to mobilize resources to fight COVID-19. As such, we included measured national income for each study in our analysis; specifically, national income is measured as the gross domestic product (GDP) subtracting capital depreciation and adding net foreign income, using data from the World Inequality Database [19].

### Statistical Analyses

#### Marginal analysis

Due to the low number of observations (n = 39 school outbreaks), we conducted an initial analysis to test combinations of the DAG covariates described above. The goal was to examine the relationship between the SAR and only one group of variables at a time and then include the most relevant predictors into the final model based on this analysis. For each test, we used a logistic regression model of the following form:

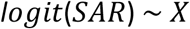

where logit is the log-odds (i.e, 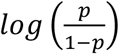, with *p* as the probability of event) and SAR represents the SAR as reported from each of the 39 school outbreaks. *X* is one of the combinations of variables we examined as follows:

1. School type
2. Classroom size, adjusting for national income, seasonal changes, and cultural climate
3. Preventive measures, adjusting for seasonal changes and cultural climate
4. Seasonal changes
5. Community transmission (weekly death rate per 100,000 or weekly case rate per 10,000, i.e., only one measure is included, because these two measures are highly correlated), adjusting for cultural climate, population immunity and study period
6. Prior population immunity, adjusting for cultural climate
7. National Income

As noted above in the conceptual analysis, the type of surveillance policy implemented in schools could affect the reported SAR in both directions. Thus, we included surveillance type in all models. However, as a sensitivity analysis, we also tested each model without surveillance type included. Results for both versions are reported in Fig 3.

**Figure 3.**
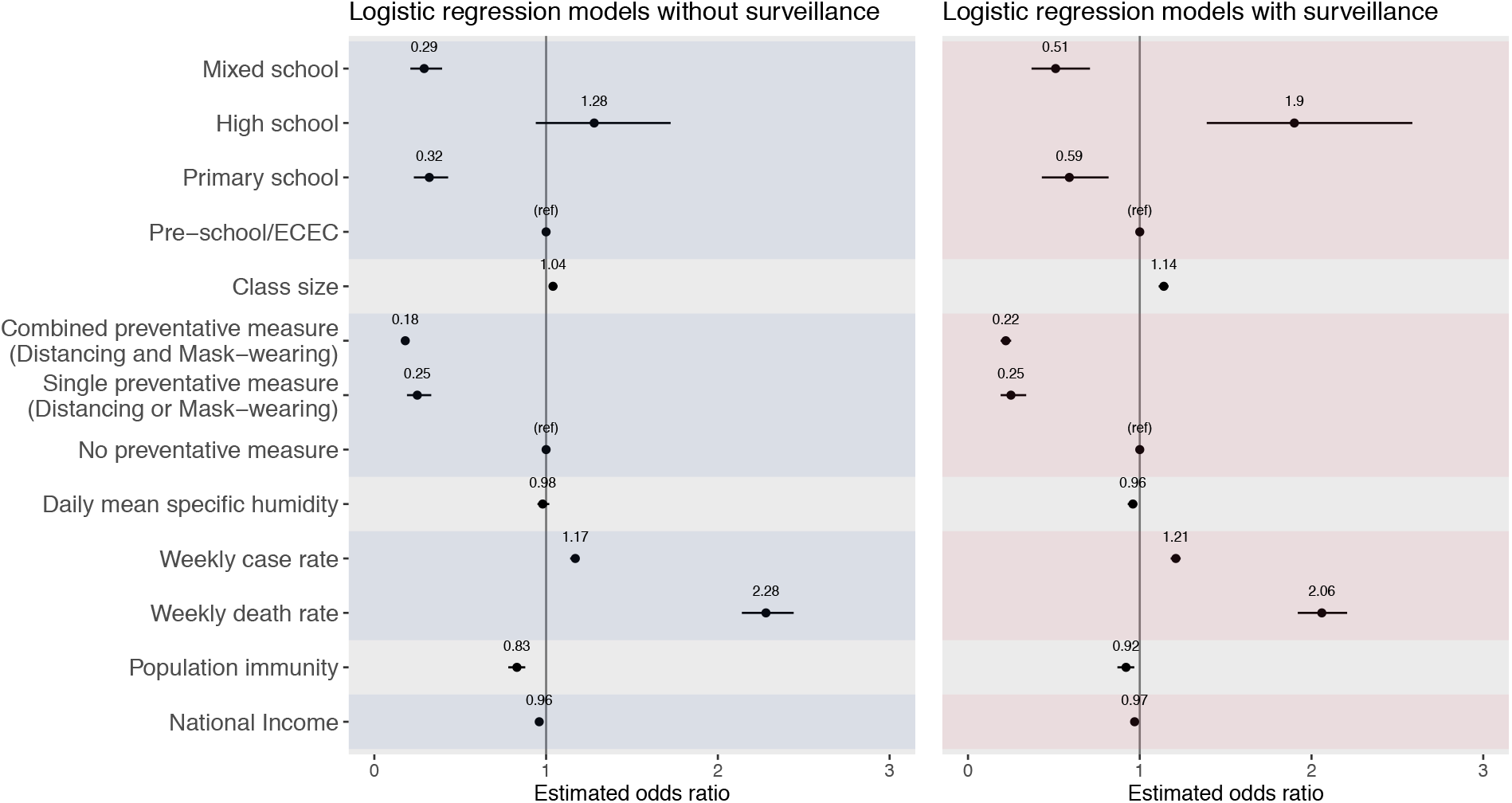
Odds ratio estimates from the marginal models. Left panel shows results from models without adjusting for surveillance and right panel shows results from corresponding models additionally adjusting for surveillance. Black points show the mean odds ratio estimates and horizontal black bars show the 95% confidence intervals. The vertical black bar in each plot indicates the null value of 1.0. Each set of models is delineated by alternating the shaded regions.

#### Multi-risk factor analysis

All seven variable groups described above were found to be associated with SAR in the marginal analysis (see Results). We thus tested models including different combinations of these variables to identify a multi-risk model that best explains the observed SAR. For all models, we included surveillance type to account for potential biases in reporting including missing asymptomatic infections, which would underestimate SAR. We also assessed for confounding between our variables of interest and SARS-CoV-2 SAR (see adjustments specified above). This procedure tested all possible combinations of significant variables identified from the marginal analysis. We then evaluated and selected the most parsimonious model with the best fit based on the Akaike information criterion (AIC; Table S1). The best performing model took the following form:

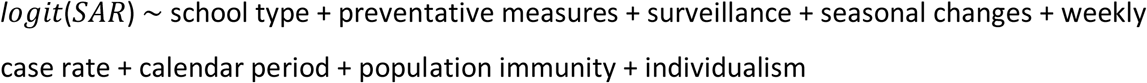

All statistical analyses were performed in RStudio, a user interface for R (R Foundation for Statistical Computing, Vienna, Austria). All models were fitted using the “glm” function from the built-in “stats” library in R.

#### Sensitivity analysis

We tested different measures of community transmission, to examine the robustness of our model results to potential biases due to variations in case-ascertainment, mortality risk, and delay in event occurrence (e.g. from infection to death) and reporting. Specifically, for the best-performing multi-risk factor model, we additionally examined three other measures in representing the intensity of community transmission, in lieu of concurrent weekly case rate: 1) weekly death rate during the study period; 2) weekly death rate during the study period plus a 1-month extension; and similarly, 3) weekly case rate during the study period plus a 1-month extension. The additional month added accounts for the time lag that occurs as death from COVID-19 may take several weeks [20].

## Results

### Summary statistics

We identified 39 reported SARS-CoV-2 outbreaks in schools, totaling 1144 secondary cases in children among 28826 contacts. These outbreaks occurred in 15 countries, spanning 4 WHO regions including the Americas, Western Pacific, European, and Eastern Mediterranean Region. Fig 1 shows the study site, school type, study period, and reported SARS-CoV-2 SAR for each included outbreak. Table 1 shows the frequencies and summary statistics for SARS-CoV-2 SAR and other variables included. While the reported SAR ranged from 0 to 100%, the majority of schools reported very low SAR (median: 2%, interquartile range: 0 – 12%). Roughly even proportion of different school types were included: 5 (13%) were pre-schools, 8 (21%) were primary schools, 11 (28%) were high schools, and 15 (38%) were mixed schools. The majority of schools tested all contacts of the index cases (25/39, or 64%); and the majority required at least one preventive measure (28/39, or 72%).

**Table 1.** Characteristics of school outbreaks and related risk factors.

| Study Characteristics | n (%), N = 39 in total |
| --- | --- |
| Calendar Period |  |
| Before July 2020 | 21 (54%) |
| After July 2020 | 18 (46%) |
| Surveillance |  |
| Symptomatic or some asymptomatic | 14 (36%) |
| All contacts | 25 (64%) |
| School type |  |
| Pre-school/ECEC | 5 (13%) |
| Primary school | 8 (21%) |
| High school | 11 (28%) |
| Mixed school | 15 (38%) |
| Preventive Measure |  |
| No preventative measure | 11 (28%) |
| Single preventative measure<br>(Distancing or Mask-wearing) | 11 (28%) |
| Combined preventative measure<br>(Distancing and Mask-wearing) | 17 (44%) |
| Higher individualism (>76) | 18 (46%) |
|  | Median (IQR) |
| Secondary attack rate | 0.02 (0.00, 0.12) |
| Population immunity rate<br>(per 100 people) | 0.36 (0.03, 1.10) |
| Weekly case rate (per 10,000 people) | 0.30 (0.03, 0.98) |
| Weekly death rate (per 100,000 people) | 0.25 (0.03, 1.79) |
| Daily mean specific humidity (g/kg) | 6.73 (4.14, 8.41) |
| National Income (thousand £) | 41 (37, 53) |
| Average class size | 20.1 (19.8, 24.0) |

### Marginal analysis

The marginal analysis with or without adjusting for surveillance generated similar estimates (Figure 3). Thus, below we present results adjusting for surveillance. This analysis identified several associating factors that are directly related to schools, including school type, class size, preventive measures, and seasonal changes. For school type, compared to pre-schools, being in primary schools (aOR: 0.59, 95% CI: 0.43 – 0.82) or mixed schools (aOR: 0.51, 95% CI: 0.37 – 0.71) was associated with a lower risk of SARS-CoV-2 infection whereas being in high schools (aOR: 1.9, 95% CI: 1.39 – 2.59) was associated with a higher risk of SARS-CoV-2 infection. For the school physical setting measure, each 1-person increase in the national average class size was associated with an increased risk of contracting SARS-CoV-2 in schools (aOR: 1.14, 95% CI: 1.11 – 1.17). Single (distancing or masking) and combined preventive measure (distancing and masking) were both associated with a lower SAR in schools, with an aOR of 0.25 (95% CI: 0.19 – 0.34) and 0.22 (95% CI: 0.19 – 0.25), respectively. For disease seasonality, which could affect the transmission in schools and the community in general, each 1 g/kg increase in specific humidity was associated with a decreased risk of contracting SARS-CoV-2 in schools (aOR: 0.96, 95% CI: 0.93, 0.99).

In addition, the marginal analysis also identified several associating factors, indirectly related to schools via the community/population. For the intensity of community transmission, both higher COVID-19 case rate and death rate in the community were associated with an increased risk of contracting SARS-CoV-2 in schools (aOR = 1.21, 95% CI: 1.18 – 1.24, for cases per 10,000 people per week; and aOR = 2.06, 95% CI: 1.92 – 2.21, for deaths per 100,000 people per week). Individualism was included in four models based on the conceptual analysis (see Methods and Fig 2); all four models showed that higher level of individualism was associated with an increased risk (mean aOR ranged from 1.3 to 2.8 and all 95% CI had a lower bound >1). Conversely, higher prior population immunity (using cumulative case rate per 100 people as a proxy, aOR: 0.92, 95% CI: 0.87 – 0.97) and higher national income (aOR: 0.97, 95% CI: 0.96 – 0.98, per 1000 pound) were associated with a reduced risk.

### Multi-risk factor analysis

Among all models tested (Table S1), the best-performing model with the lowest AIC included six key groups of risk factors, namely, school type, preventive measures, seasonality, intensity of community transmission, population immunity, and individualism, adjusting for surveillance. Overall, these risk factors in combination were able to explain 60.6% of the variance in the reported SARS-CoV-2 SAR (McFadden’s pseudo R^2^ = 0.606; Fig 4) [21, 22]. The estimated adjusted ORs (aORs) for each risk factor are shown in Table 2 and Fig 5. The sensitivity analysis shows consistent estimates across models using different measures of community transmission (Table S2).

**Figure 4.**
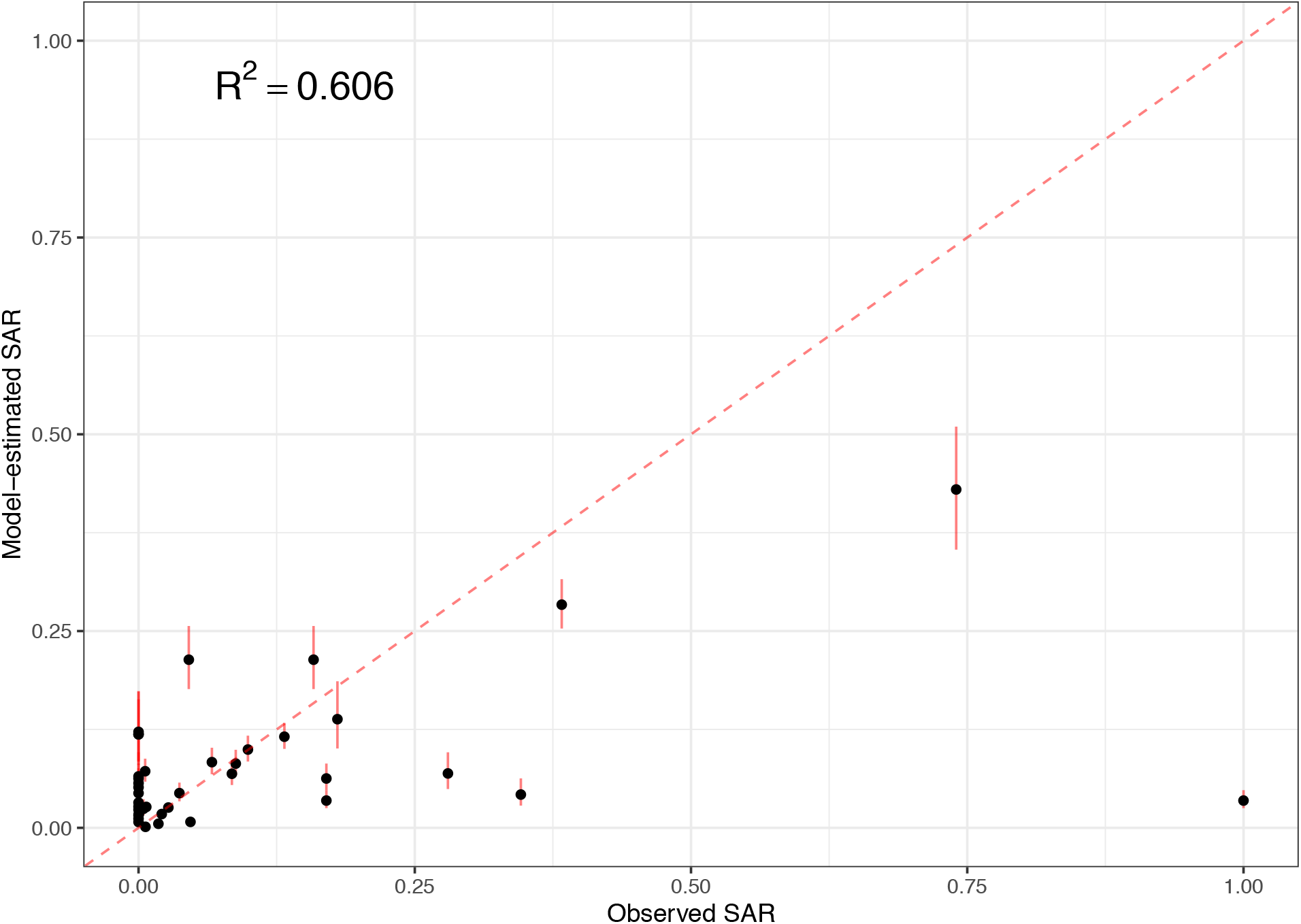
Model fit of the best-performing multi-risk model. Black dots show the fitted SAR for each study (y-axis), compared to the observed SAR (x-axis); the red bars around each point show the 95% confidence intervals of model-estimates. The McFadden’s pseudo-R^2^ is computed using Eqn. 30 in [22].

**Table 2.** Results of the best-fit multi-risk factor model for the identification of factors associated with SARS-CoV-2 SAR in schools. Adjusted odds ratio (aOR) estimates and 95% confidence intervals are given from the logistic regression model including surveillance to control for differences in testing school clusters, school type due to inconsistent reporting of age groups in the literature, number of preventative measures implemented in schools, and characteristics of the study sites (i.e., level of individualism, daily mean specific humidity, population immunity and weekly case rates per 10,000).

| Variable | aOR (95% CI) |
| --- | --- |
| School type |  |
| Mixed school | 0.24 (0.16, 0.36) |
| High school | 1.3 (0.9, 1.88) |
| Primary school | 0.35 (0.23, 0.54) |
| Pre-school/ECEC | reference |
| Preventive measures |  |
| Combined preventative measure<br>(Distancing and Mask-wearing) | 0.22 (0.17, 0.29) |
| Single preventative measure<br>(Distancing or Mask-wearing) | 0.22 (0.15, 0.34) |
| No preventative measure | reference |
| Seasonal changes |  |
| Daily mean specific humidity | 0.96 (0.93, 1) |
| Community transmission |  |
| Weekly case rate | 1.2 (1.17, 1.24) |
| Population immunity |  |
| Population immunity | 0.28 (0.22, 0.35) |
| Individualism |  |
| Higher individualism | 1.72 (1.19, 2.5) |
| Lower individualism | reference |
| Surveillance |  |
| All contacts | 0.59 (0.5, 0.7) |
| Symptomatic or some asymptomatic | reference |
| Calendar Period |  |
| After July 2020 | 1.93 (1.45, 2.59) |
| Before July 2020 | reference |

**Figure 5.**
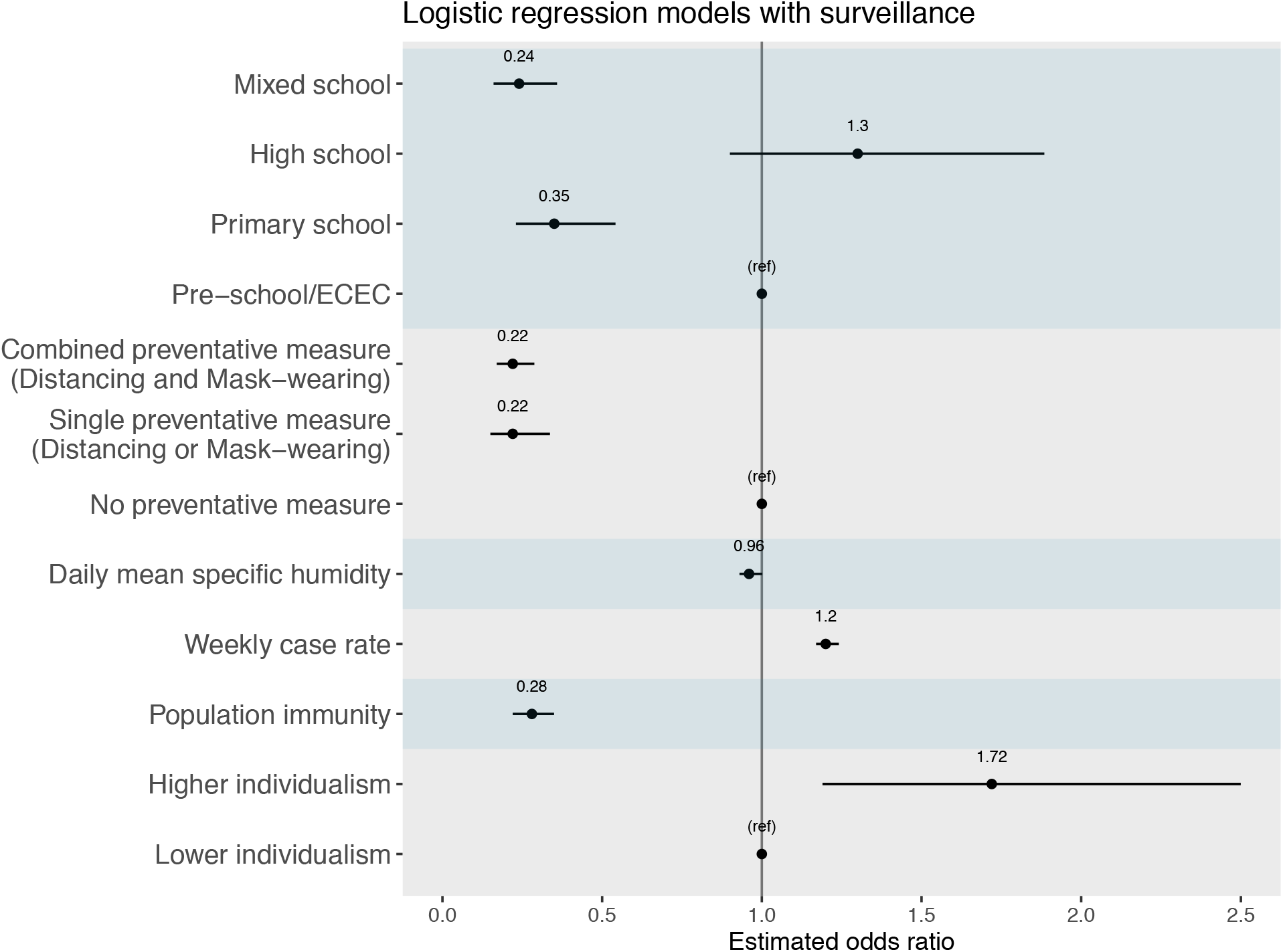
Odds ratio estimates from the best-performing multi-risk model. Black points show the mean odds ratio estimates and horizontal black bars show the 95% confidence intervals. The vertical black bar indicates the null value of 1.0. Each variable type is delineated by the shaded regions.

Consistent with the marginal analysis, the best-fit multi-risk factor model showed that higher COVID-19 case rate in the community (aOR: 1.2, 95% CI: 1.17 – 1.24; for 1 additional case reported among 10,000 people each week) and higher level of individualism (aOR: 1.72, 95% CI: 1.19 – 2.5; above vs. below the median) were associated with an increased risk of SARS-CoV-2 infection in schools. Conversely, both single (aOR: 0.22, 95% CI: 0.15 – 0.34) and combined (aOR: 0.22, 95% CI: 0.17 – 0.29) preventive measure were associated a reduced risk of SARS-CoV-2 infection in schools. In addition, both testing all contacts (aOR: 0.59, 95%: 0.50 – 0.70; vs. only testing the symptomatic or some asymptomatic) and higher population immunity (aOR: 0.28, 95% CI: 0.22 – 0.35, using cumulative case rate per 100 people as a proxy) were also associated a reduced risk of SARS-CoV-2 infection in schools. Compared to students in pre-schools, the aOR was 0.24 (95% CI: 0.16 – 0.36) for students in primary schools, 1.3 (95% CI: 0.9 –1.88) for students in high schools, and 0.24 (95% CI: 0.16 – 0.36) for students in mixed schools.

## Discussion

Leveraging available data on multiple reported SARS-CoV-2 outbreaks in schools and potential risk factors, we have examined main factors associated with the risk of SARS-CoV-2 transmission in schools. Our analyses suggest that SARS-CoV-2 SAR in schools was associated with both preventative measures in schools and population factors including the level of community transmission, individualism, and population immunity, once adjusted for surveillance and school type.

Foremost, we identified several population or community factors to be highly associated with SARS-CoV-2 transmission in schools, all of which point to the importance of communal efforts to collectively reduce the risk of transmission and protect children in schools. In particular, all models (in both the marginal analysis and the multi-risk factor analysis) consistently showed that higher level of individualism of the population was associated with higher SARs in schools. This finding is consistent with a recent study linking collectivism (vs. individualism) to usage of preventive measures like mask use during the COVID-19 pandemic [23]. Along similar lines, the models associated higher transmission in the community with higher SARs in schools, suggesting the potential community-to-school importation of cases and subsequent risk of outbreak in schools. As such, care must be applied when reopening or operating schools in areas with high levels of community transmission. In addition, reversing some original fears about school-to-community SARS-CoV-2 transmission, it is likely that the community transmission drives outbreaks in school, not the reverse. Further, the models showed that higher population immunity, which could lower transmission overall, was associated with lower SARs in schools. With the availability of COVID-19 vaccines, predominantly to adults at present, it is paramount that all eligible adults get vaccinated promptly to lower the risk of transmission in the community and in turn to provide indirect protection to children via the increased population immunity.

Our models estimated a substantial transmission reduction in schools when both distancing and mask-wearing were required (aOR: 0.22, 95% CI: 0.17 – 0.29) or when distancing alone was required [note the aOR for either measure alone was 0.22 (95% CI: 0.15 – 0.34), with the majority of schools in this category (8 of 11) requiring distancing alone]. Thus, both estimates indicate the importance of distancing. This finding is likely a combined outcome of reduced number of contacts and reduced short-range transmission when social distancing policies were followed. Maintaining distance has necessitated fewer people in a room at the same time, leading to fewer contacts. In addition, the increased personal space in classroom enables students to avoid the likely higher viral concentration within short-range of the emitter (either via aerosols, droplets, or in combination) when far apart. Nevertheless, it is important to note that social distancing measures may be more difficult to achieve fully in disadvantaged communities (often of color) with underfunded and overcrowded schools [24, 25]. Furthermore, racially motivated structural factors prevent these disadvantaged communities from practicing social distancing policies outside of the school. For example, these communities tend to make up most of essential workers and thus have higher rates of transmission in their community [26], increasing potential introduction of infections into schools.

In comparison to pre-schools, students in both primary and mixed-level schools had a lower risk of SARS-CoV-2 infection whereas those in high schools had a higher risk. This finding is consistent with previous studies indicating the likely lower susceptibility to SARS-CoV-2 infection and transmissibility among young children [4, 5]. In addition, it is also likely in part due to the greater ability of older children to follow directions regarding preventive measures but with less compliance among high school students. Overall, these findings support flexible reopening policies for different levels of schools given the risk differences.

When schools test all contacts of the index case, this effectively functions as a control measure in that more cases will be detected and is more likely to result in a school closure. This may explain why the OR estimate for all contacts surveillance is protective, as the outbreak was identified and able to be contained before the asymptomatic individuals were able to infect others.

This study has several limitations. First, all school outbreaks included in this analysis occurred prior to the emergence and widespread circulation of the more transmissible SARS-CoV-2 variants of concern (e.g., the delta variant). We are thus unable to estimate variant-specific impacts. Nonetheless, even though the magnitude of impact may alter somewhat due to changes in circulating SARS-CoV-2 variants, the identified risk factors and their relative importance to school transmission likely would still hold given the robust risk mechanisms. Second, we were unable to estimate the impact of distancing and mask-wearing separately, due to the small sample size of schools that required masking alone (n = 3). Third, due to a lack of detailed information for each specific school setting, we used proxy measures in the analyses (e.g., class size at the national level was used rather than for each reporting school), which may have limited the ability of the models to identify the association of these factors with SARS-CoV-2 transmission risk. Similarly, due to the lack of data, we were not able to examine other key factors such as ventilation in classrooms, social economic status of individual students and their households, and potential differences in susceptibility and transmissibility by age group. Future work with comprehensive study designs and data collection is warranted to provide further insights into how infections, not limited to SARS-CoV-2, spread in schools and the broad, bi-directional impact of school and community transmission. This would be invariable to inform better strategies to combat future infectious disease outbreaks.

## Data Availability

All data are publicly available as detailed in the paper.

## Author contributions

WY conceived and supervised the study; HY, CR, and SN conducted the literature review and data extraction, and performed the statistical analysis; HY, CR, SN, and WY interpreted results and wrote the manuscript.

## Supplemental Tables

**Table S1.**
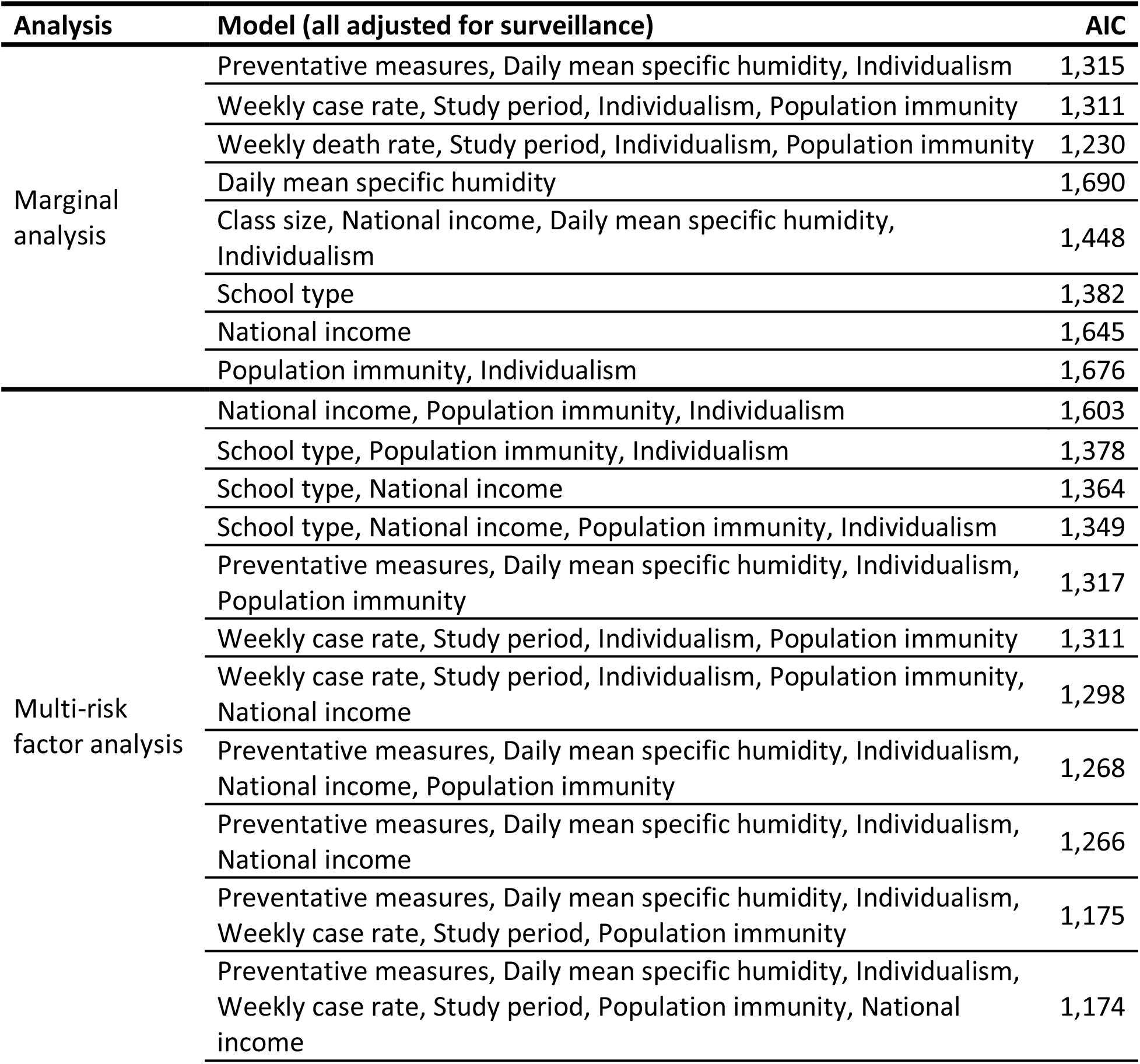
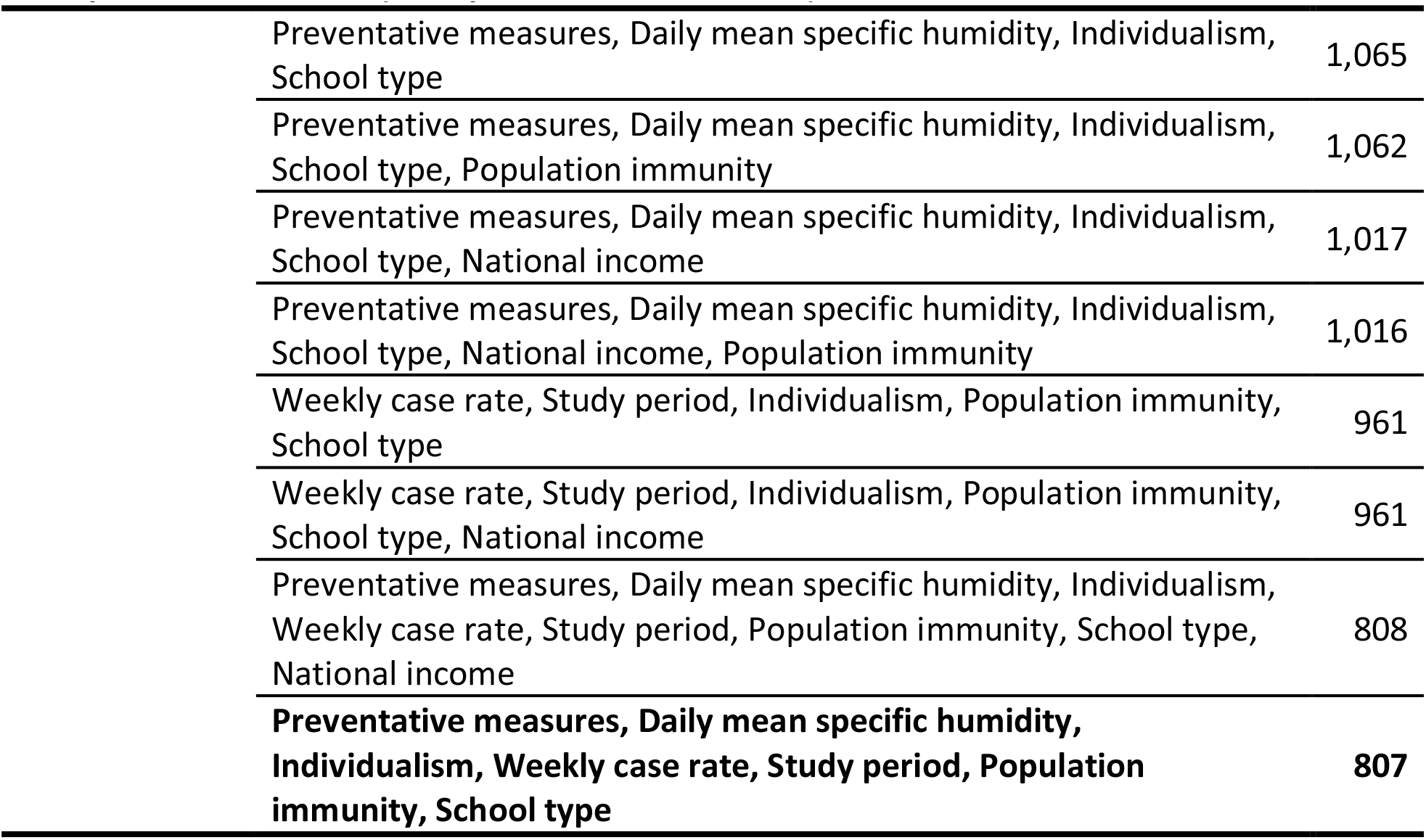
Performance of different models. In the marginal analysis, 8 models for 7 groups of risk factors were tested (note weekly case rate and weekly death rate, both representing community transmission, were tested separately in two models). In the multi-risk factor analysis, 19 additional models with all other possible combinations of 6 major groups of risk factors tested significant (i.e., when a risk group was a subset of a larger one, only the larger risk group was tested). All models adjusted for surveillance. The best-performing model with the lowest AIC is bolded.

**Table S2.** Sensitivity analysis for both cases and deaths, with and without a 1-month extension to the study time period.

| Variable |  | Without extension |  | With extension |  |
| --- | --- | --- | --- | --- | --- |
|  |  | aOR (95%CI) | AIC | aOR (95%CI) | AIC |
| case | Weekly case rate | 1.2 (1.17, 1.24) | 807.34 | 1.07 (1.05, 1.08) | 924.49 |
|  | Single preventative measure (Distancing or Mask-wearing) | 0.22 (0.15, 0.34) |  | 0.16 (0.11, 0.22) |  |
|  | Combined preventative measure (Distancing and Mask-wearing) | 0.22 (0.17, 0.29) |  | 0.21 (0.16, 0.27) |  |
|  | Primary school | 0.35 (0.23, 0.54) |  | 0.32 (0.21, 0.47) |  |
|  | High school | 1.3 (0.9, 1.88) |  | 1.01 (0.72, 1.42) |  |
|  | Mixed school | 0.24 (0.16, 0.36) |  | 0.19 (0.13, 0.27) |  |
|  | All contacts | 0.59 (0.5, 0.7) |  | 0.78 (0.67, 0.91) |  |
|  | After July 2020 | 1.93 (1.45, 2.59) |  | 2.13 (1.59, 2.84) |  |
|  | Population immunity | 0.28 (0.22, 0.35) |  | 0.61 (0.54, 0.69) |  |
|  | High individualism (>76) | 1.72 (1.19, 2.5) |  | 2.12 (1.54, 2.91) |  |
|  | Daily mean specific humidity | 0.96 (0.93, 1) |  | 0.87 (0.84, 0.91) |  |
| death | Weekly death rate | 1.73 (1.59, 1.89) | 843.64 | 1.27 (1.18, 1.37) | 940.31 |
|  | Single preventative measure (Distancing or Mask-wearing) | 0.2 (0.14, 0.28) |  | 0.19 (0.13, 0.27) |  |
|  | Combined preventative measure (Distancing and Mask-wearing) | 0.18 (0.14, 0.23) |  | 0.2 (0.15, 0.26) |  |
|  | Primary school | 0.39 (0.25, 0.58) |  | 0.54 (0.35, 0.82) |  |
|  | High school | 0.89 (0.62, 1.27) |  | 1.62 (1.13, 2.34) |  |
|  | Mixed school | 0.28 (0.19, 0.41) |  | 0.36 (0.24, 0.53) |  |
|  | All contacts | 1.14 (0.95, 1.37) |  | 0.86 (0.74, 1.02) |  |
|  | After July 2020 | 2.48 (1.9, 3.24) |  | 2.55 (1.94, 3.35) |  |
|  | Population immunity | 0.7 (0.64, 0.77) |  | 0.78 (0.72, 0.85) |  |
|  | High individualism (>76) | 1.95 (1.42, 2.67) |  | 2.19 (1.6, 3) |  |
|  | Daily mean specific humidity | 0.99 (0.95, 1.03) |  | 0.95 (0.91, 0.99) |  |

